# LCK and HOMER1 gene expression-based classifier distinguishes early mycosis fungoides from eczema and psoriasis

**DOI:** 10.1101/2025.10.18.25338021

**Authors:** Martin Meinel, Nora Langreder, Andrea Schmitt, Kristin Technau-Hafsi, Christina Hillig, Sophie Roenneberg, Adriana Bernklauová, Jan Říčař, Denisa Kacerovská, Marion Wobser, Caroline Mann, Beate Weidenthaler-Barth, Yalda Ghoreishi, Reem El Bahtimi, Leonidas Marinos, Evangelia Papadavid, Marie Charlotte Schuppe, Christina Mitteldorf, Werner Kempf, Emmanuella Guenova, Kilian Eyerich, Michael P. Menden, Stefanie Eyerich, Natalie Garzorz-Stark

## Abstract

Mycosis fungoides (MF) is a rare cutaneous T cell lymphoma that, in its early stages, can closely mimic eczema and psoriasis in both clinical appearance and histopathologic features, leading to frequent misdiagnosis, inappropriate treatment, and delayed care. Reliable adjunctive biomarkers are lacking, underscoring the need for improved diagnostic strategies. We developed a biomarker discovery framework based on bulk RNA sequencing of skin biopsies (19 MF, 112 psoriasis, 105 eczema), which identified seven candidate diagnostic genes. RT-PCR analysis in FFPE tissue specimens from 65 MF and 101 eczema/psoriasis samples verified *LCK* and *HOMER1* as robust discriminator genes. A logistic regression model based on *LCK* and *HOMER1* gene expression differentiated MF from psoriasis and eczema with 91% sensitivity and 94% specificity. Independent validation on 7 additional international cohorts (MF n=58, eczema/psoriasis n=55) confirmed robust performance. Spatial and single-cell transcriptomic analyses revealed biological underpinnings of classifier accuracy: *LCK* was enriched in malignant and specific T cell subsets in MF, whereas *HOMER1* was confined to keratinocytes in eczema and psoriasis but nearly absent in MF. Case studies demonstrated that the classifier identified MF in routine biopsies earlier than histopathology.

This molecular diagnostic approach enables earlier and more reliable distinction of MF from common inflammatory dermatoses, offering a clinically applicable tool to reduce diagnostic uncertainty, accelerate appropriate treatment, and might improve patient outcomes.

**One Sentence Summary:** A two-gene classifier reliably distinguishes mycosis fungoides from eczema and psoriasis, enabling early and accurate diagnosis.

## INTRODUCTION

Mycosis fungoides (MF) is the most common subtype of cutaneous T cell lymphoma (CTCL) and accounts for more than 60% of all primary cutaneous lymphomas (*1, 2*). The condition is characterized by infiltration of malignant T cells into the skin and manifests with a wide clinical spectrum, ranging from erythematous patches and plaques to advanced tumor-stage disease. In the early stage, when 80% of patients first present, MF often mimics benign dermatoses such as eczema or psoriasis. Due to this similarity, MF has earned the designation of “great simulator” or a “chameleon of dermatology” (*3, 4*). This overlap makes early diagnosis particularly difficult, as often neither clinical evaluation nor histopathology reliably distinguishes MF from benign inflammatory skin diseases (*5*). Conventional T cell receptor clonality testing used as diagnostic support lacks reliability, showing polyclonality in MF and *vice versa* monoclonality in benign conditions such as lichen planus or pityriasis lichenoides (*6, 7*). The consequences of delayed recognition are profound. Data from the international PROCLIPI registry showed a median delay of 36 months between symptom onset and diagnosis, with 85% of patients affected (*8*). During this time, patients might receive inappropriate treatment with biologics or immunomodulators intended for inflammatory skin diseases, which may worsen outcomes or even trigger development of MF (*9–12*). Moreover, delayed diagnosis may facilitate progression to advanced stages that are associated with substantial deterioration in quality of life, including disfiguring and painful skin lesions, severe pruritus, and psychosocial distress (*13*). At these stages, patients often require antineoplastic systemic treatment (*14*), driving healthcare expenditures to a level approximately four times higher than for early-stage lymphomas (*15*). Thus, early and accurate diagnosis is essential to enable appropriate patient management, inform therapeutic decisions, and avoid unnecessary or ineffective interventions, thereby potentially reducing patient burden and healthcare costs.

Recent advances in molecular profiling, including gene expression and miRNA classifiers, have identified promising biomarkers such as TOX, PDCD1, TRAF1, and others (*16–20*). While these studies highlight the potential of molecular diagnostics to overcome current limitations, no robust biomarker has yet been established for routine clinical use. This unmet need motivates the development of reliable, clinically applicable molecular tools to improve the early detection of MF and prevent harmful misdiagnosis. Eczema and psoriasis may be diagnostically indistinct from MF on clinical and histopathologic grounds; however, MF remains uncommon (∼1 per 100,000), in contrast to much higher prevalence of eczema and psoriasis (≥5% of adults in developed countries; >300 million worldwide (*21–23*). This pronounced discrepancy in prevalence poses a major challenge for molecular studies, as rare diseases like MF are often underrepresented in biomarker discovery cohorts. To address this limitation, we curated cohorts reflecting real-world diagnostic ambiguity and sufficient MF representation, and developed **<u>S</u>**parse biom**<u>A</u>**rkers **<u>F</u>**or r**<u>AR</u>**e d**<u>I</u>**seases (SAFARI), a biomarker discovery framework designed to identify sparse yet robust gene signatures that remain reliable even in the context of small and imbalanced sample sets. Applying SAFARI to our bulk RNA-seq discovery cohort, we identified *LCK* and *HOMER1* as powerful markers for distinguishing early-stage MF from psoriasis and eczema. Their diagnostic utility was independently demonstrated using quantitative PCR in formalin-fixed paraffin embedded (FFPE) skin biopsies of routine histopathology. Validation in seven independent international cohorts demonstrated reproducibility and generalizability across multiple clinical sites. Moreover, in sequential biopsies from patients at a later timepoint diagnosed with MF the classifier detected disease earlier than conventional methods, highlighting its potential as a critical tool for early diagnosis. These findings establish a molecular framework for earlier and more accurate diagnosis of MF, with the potential to adapt therapeutic strategies and improve patient outcomes.

## RESULTS

### Clinical, histological, and broad transcriptomic overlap in mycosis fungoides is resolved by unique expression signatures

Clinical and histopathologic similarity to inflammation complicates the diagnosis of early MF, as lesions often resemble common inflammatory dermatoses such as eczema and psoriasis. Representative cases illustrate this challenge (Fig. 1A): Eczema typically shows spongiosis and serum crust, sometimes with acanthosis and dermal lymphocytic infiltrates, whereas psoriasis is characterized by psoriasiform epidermal hyperplasia, parakeratosis often with inclusions of neutrophils, hypogranulosis, elongated, tortuous, and dilated capillaries in dermal papillae, and mixed lymphocytic–neutrophilic infiltrates. Both patterns, however, can also occur in MF, which presents with heterogeneous features ranging from eczematous to psoriasiform changes, including parakeratosis, acanthosis, spongiosis, and dermal infiltrates of lymphocytes and neutrophils. To explore whether molecular profiling would allow for separation of MF from psoriasis and eczema, we performed bulk RNA sequencing of lesional and non-lesional skin biopsies from 19 MF, 105 eczema, and 112 psoriasis patients (“discovery cohort”) (Fig. 1B, Table S1). MF was underrepresented in the discovery cohort and as such reflects the natural incidence of this rare disease in comparison to eczema and psoriasis. Dimensionality reduction by UMAP revealed partially distinct clustering of MF samples compared with eczema and psoriasis, suggesting disease-specific transcriptional signatures (Fig. 1C). We next thought to identify MF specific genes and performed comparative gene expression analysis. A panel of genes significantly upregulated in MF compared with psoriasis and eczema was identified including immune-related genes such as *CXCL13*, *GNLY*, and *SH2D1A*, while inflammatory mediators including *S100A2*, *IL20*, and *CXCL8* were significantly downregulated in MF compared with psoriasis and eczema (Fig. 1D). Functional enrichment of genes differentially expressed genes in MF versus psoriasis and eczema revealed pathways central to T cell biology such as immune synapse organization, immunoregulatory interactions between lymphoid and non-lymphoid cells as well as chemokine receptor signaling, consistent with a T cell driven malignancy shaped by its tumor microenvironment (Fig. 1E). Together, these findings demonstrate that, although early MF shares considerable clinical, histological, and transcriptomic overlap with eczema and psoriasis, specific genes and pathways may separate MF from inflammatory skin diseases.

**Fig. 1.**
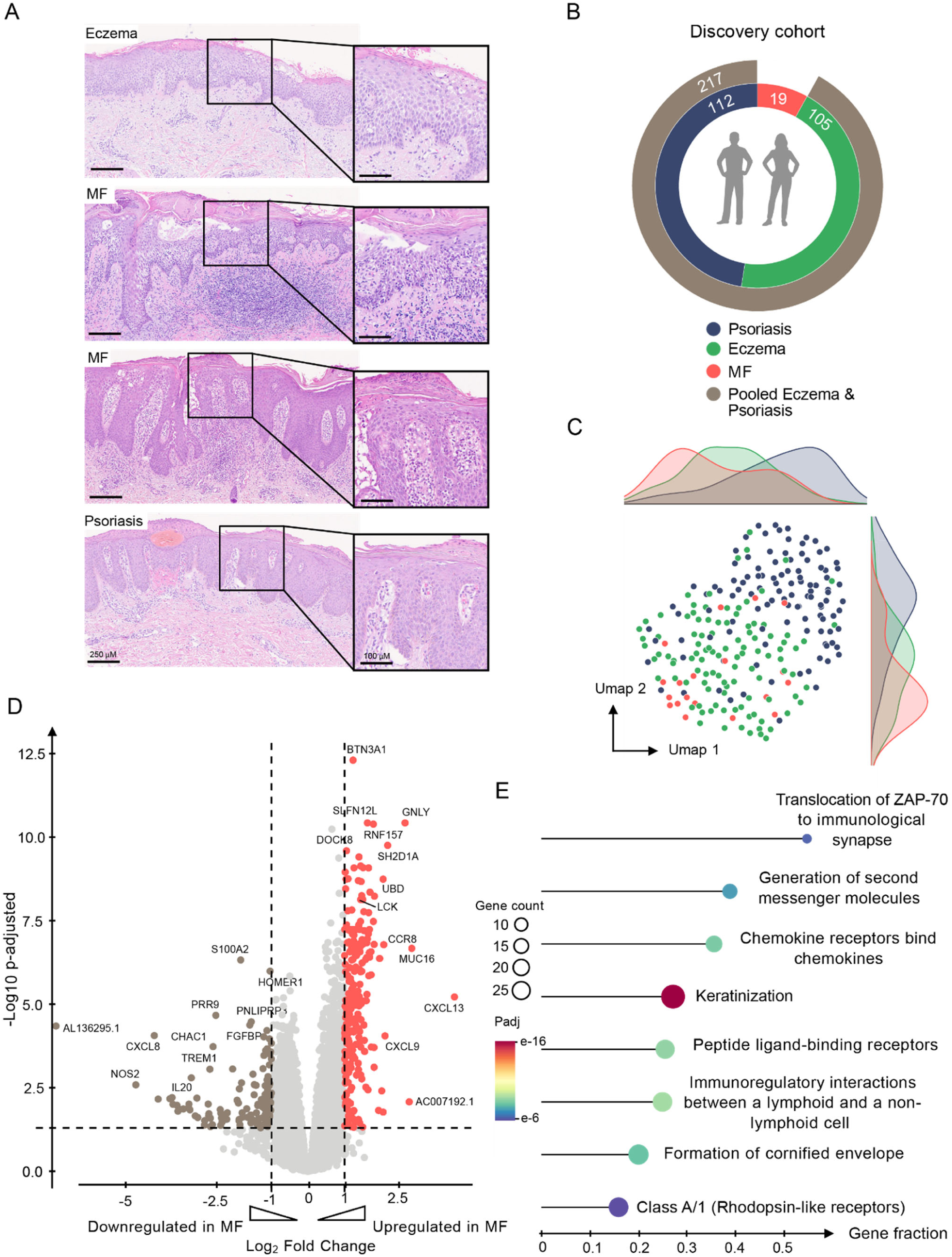
MF presents a specific molecular signature that discriminates it from inflammatory diseases. (**A**) Histology picture of eczema (top), MF (middle) and psoriasis (bottom). (**B**) Overview of the discovery cohort. (**C**) Clustering of MF (red dots) as well as eczema (green dots) and psoriasis (blue dots) samples in the UMAP space. (**D**) Significantly regulated genes in MF in comparison to eczema/psoriasis. (**E**) Over-representation analysis (ORA) of significantly regulated MF genes and their associated pathways. MF: Mycosis fungoides; ORA: Over-representation analysis. Clinical images in A available upon request from the corresponding author.

### Iterative feature selection reveals a sparse and reproducible biomarker signature for MF

The clinical translation of biomarkers requires a sparse panel of genes with robust and reproducible expression across technical and biological contexts, a challenge that is particularly pronounced in rare diseases such as MF. To meet this need, we developed the **SAFARI** framework, inspired by principles from Chen et al. (*24*) which integrates filter-, wrapper-, and embedding-based methods for biomarker discovery in medical data (Fig. 2). Since gene-expression based biomarkers distinguishing eczema from psoriasis are already well established (*25*), we pooled eczema and psoriasis samples in the following analyses, focusing on the more clinically relevant task of differentiating early MF from benign inflammatory dermatoses. SAFARI combines univariate and multivariate approaches within a bootstrapping scheme to maximize sample utility and ensure stability of results (Fig. 2A). Applied to bulk RNA seq data from our discovery cohort of 19 MF, 105 eczema, and 112 psoriasis samples, SAFARI repeatedly (200 iterations) allocated subsets of patients for univariate differential gene expression analysis to identify individually discriminatory genes, while excluding transcripts with low expression levels to enhance reproducibility in downstream qPCR validation (Fig. 2B, C). The remaining samples were then subjected to multivariate analysis, where candidate genes were ranked according to their combined discriminatory performance using sequential forward feature selection with logistic regression (Fig. 2D). To consolidate findings, RobustRank aggregation (*26*) was applied across all iterations, prioritizing genes consistently selected as strong classifiers (Fig. 2E). False discovery rates were controlled using a two-step correction accounting for both the number of genes tested per run and the number of bootstrap repeats. Eventually, we identified seven genes *HOMER1*, *LCK*, *RNF213*, *ZC3H12D*, *SERBP1*, *BTN3A1*, and *PNLIPRP3* as candidate biomarkers showing the most significant difference between MF and pooled eczema/psoriasis (Fig. 2F).

**Fig. 2.**
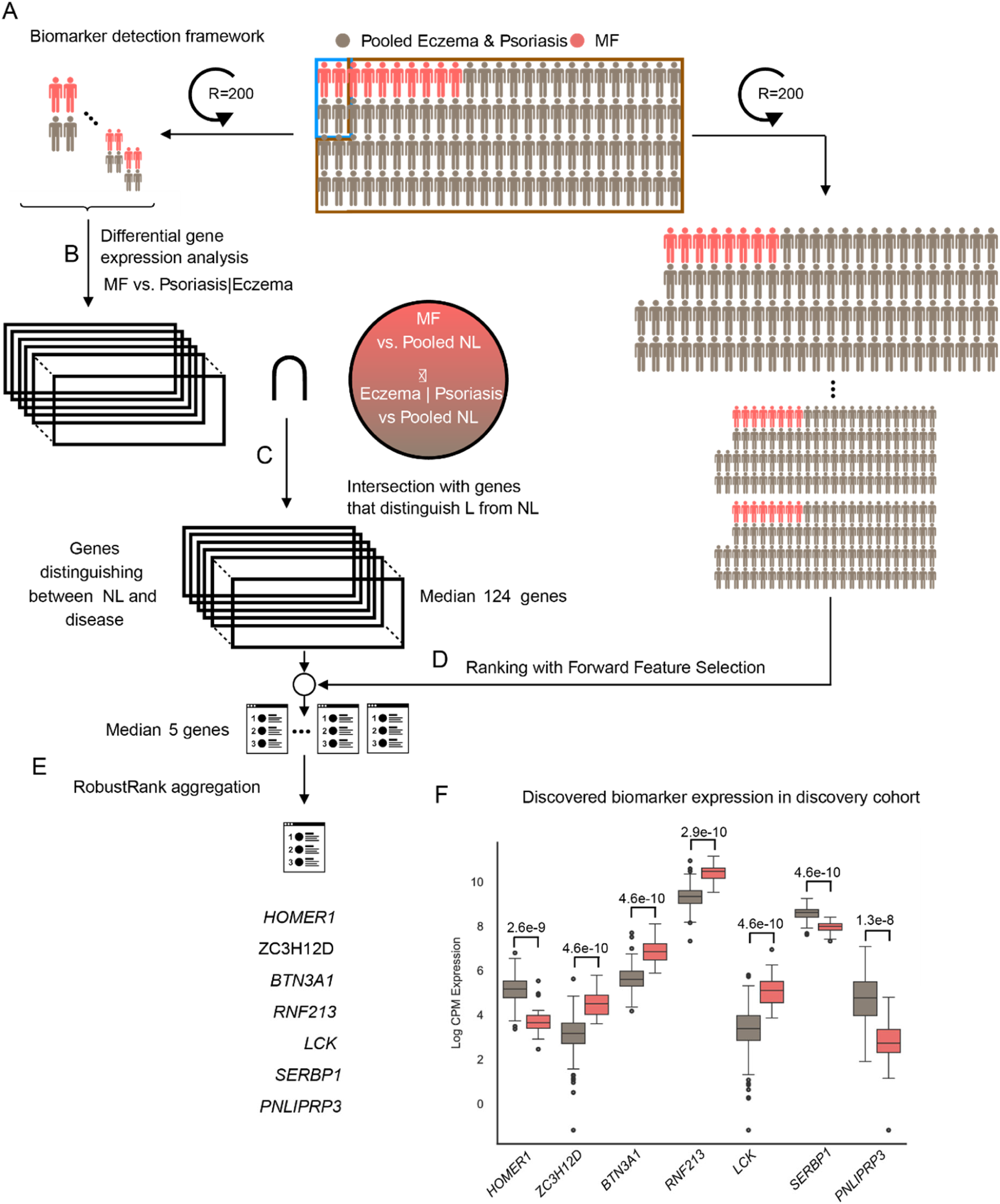
SAFARI identifies biomarkers for MF diagnosis (A) Overview of the SAFARI workflow. **(A)** (**B**) A small subset equally representing both conditions is sampled from the discovery cohort to extract differentially expressed genes between the two conditions of interest. (**C**) These genes are intersected with genes differentially expressed between lesional MF and eczema and psoriasis, and pooled non-lesional samples. (**D**) The remaining samples, not used for the differential gene expression analysis, are leveraged to rank the intersected genes by their complimentary discriminative power using sequential forward feature selection. The entire process is repeated for two 200 runs in a bootstrapping fashion enforcing robustness by providing variety in the subset compositions and consequently in the features. (**E**) Finally, Robustrank aggregation is implemented to reveal the biomarkers significantly ranked on top the 200 ranked feature sets resulting from the bootstrapping. (**F**) Expression of selected markers in the discovery cohort. NL= no-lesional, L = lesional.

### Validation in FFPE samples identifies *LCK* and *HOMER1* as robust discriminators of MF

To validate the discriminatory capacity of the biomarker panel identified by SAFARI, we established an independent development cohort consisting of the two subcohorts DE1 and DE2 comprising in total 65 MF samples, 25 eczema samples, 37 psoriasis samples and 39 samples showing both features of psoriasis and eczema (EPV = eczema-psoriasis overlap variants) (Fig. 3A, Fig. S1A, Table S1). Marker expression was quantified by qPCR from FFPE tissue, and while *PNLIPRP3* gene expression values did not pass quality control due to high standard deviation the six candidates *HOMER1*, *LCK*, *RNF213*, *ZC3H12D*, *SERBP1*, *BTN3A1* showed significant expression differences between MF and pooled eczema/psoriasis samples but also when eczema, psoriasis and EPV were examined separately across both cohorts (Fig. S1B and C). Among these, *LCK* and *HOMER1* provided the strongest separation (Fig. 3B). Mapping patient samples according to *LCK* and *HOMER1* expression levels revealed a clear separation of MF from pooled eczema and psoriasis samples (Fig. 3C, Fig. S1D). A logistic regression classifier based on *LCK* and *HOMER1* was developed including a step of score calibration to improve alignment of predictions with true class labels (Fig. S2A and B). In addition, a rejection interval was implemented to identify the optimal compromise between sensitivity, specificity, and rejection interval width, thus balancing diagnostic accuracy with the proportion of samples excluded and not definitely assigned to either MF or eczema/psoriasis (Fig. S2C). A robust performance with an AUC of 0.97 in receiver operating characteristic (ROC) analysis was achieved and the classifier reliably distinguished MF from pooled eczema and psoriasis samples but also when MF, eczema, psoriasis and EPV were examined separately across both cohorts (Fig. 3D, Fig. S 2D, Table S4). Four out of 15 misclassifications and in total 4.8 % of all samples fell within the predefined rejection interval (Fig. 3E, F) and overall, the model reached a sensitivity of 0.91, specificity of 0.94, and balanced accuracy of 0.93 (Fig. 3G).

**Fig. 3.**
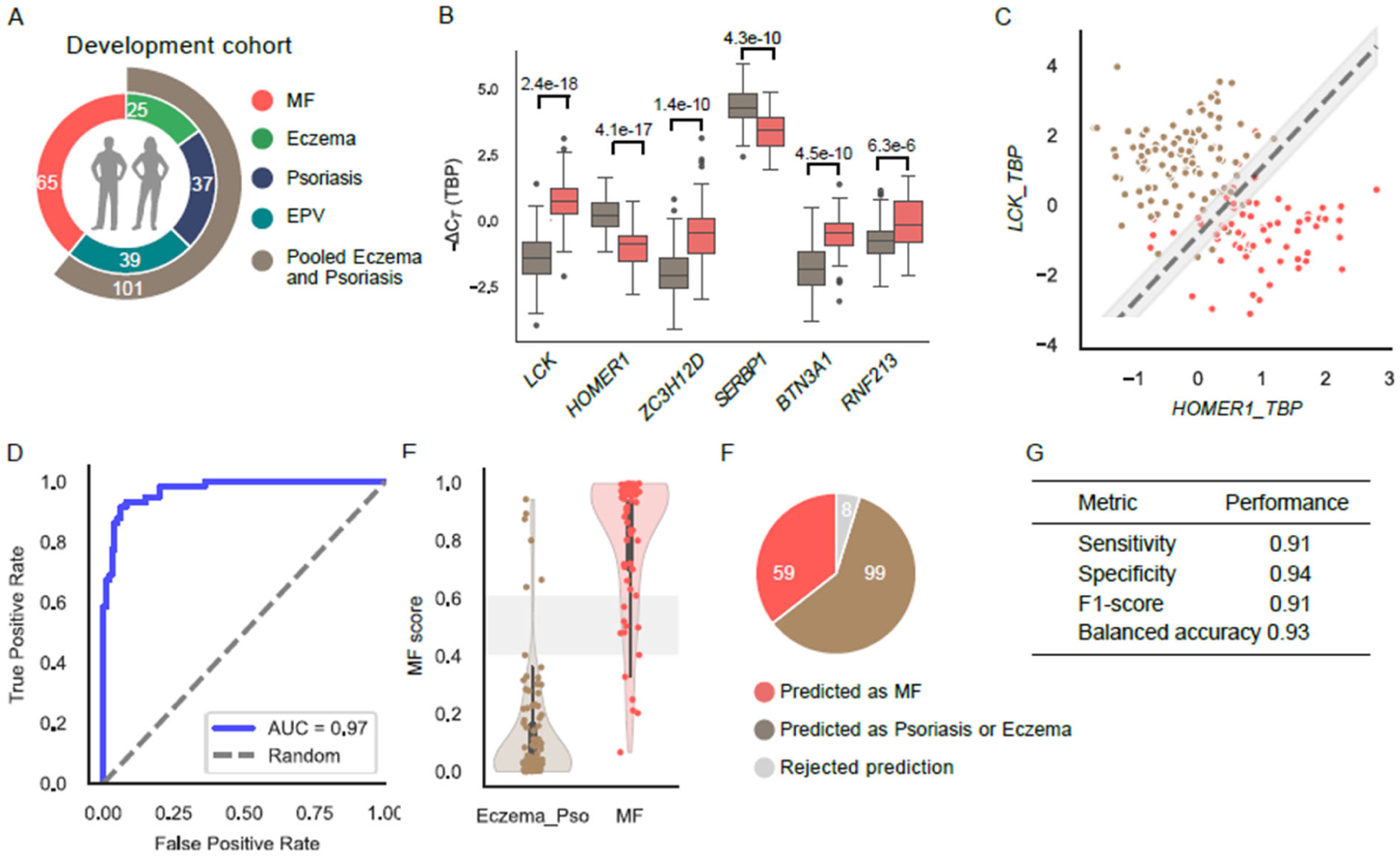
The MF classifier efficiently diagnoses MF in development cohorts. **(A)** Composition of the development cohort including MF (n=65) and pooled eczema/psoriasis (n=101) comprising eczema (n=25), psoriasis (n=37) and EPV (n=39). **(B)** Expression of selected markers by real time PCR in FFPE lesional skin samples (n=65 MF (red) and n=101 Eczema and Psoriasis samples (brown)). Shown is the delta C_T_ to the reference gene TBP. **(C)** Projection of MF and pooled eczema/psoriasis samples into LCK – HOMER1 expression space (normalized to TBP) including the rejection interval. **(D)** Receiver operating characteristics (ROC) analysis of the MF classifier consisting of LCK and HOMER1. **(E)** Scores for MF for each diagnostic group with shaded rejection interval (≤ 0.6 and ≥ 0.4). (**F**) Classification outcome distribution indicating numbers of samples predicted as MF, as eczema/psoriasis and with rejected prediction. **(G)** Performance metrics in the test cohort listing sensitivity, specificity, F1 score, and balanced accuracy (0.93). EPV = eczema-psoriasis overlap variants

### *LCK* and *HOMER1* display distinct, disease-specific spatial and cellular expression patterns underpinning classifier performance

To place the two gene-classifier in biological context and understand if the markers would relate to known processes in lymphoma biology, we next examined the spatial and cellular distribution of *LCK* and *HOMER1* within the tissue microenvironment using previously published spatial transcriptomics and single cell datasets on MF (*27, 28*), psoriasis and eczema (*29*). H&E-integrated spatial gene expression maps in lesional eczema and psoriasis hinted at strong expression of *HOMER1* in middle/upper- and basal epidermis as well as in the upper dermis, while it showed an overall lower expression in MF (Fig. S3A, B). In contrast, *LCK* was mainly expressed in the epidermal compartment of eczema and psoriasis, while in MF it could be specifically allocated to malignant CD4⁺ T cells in the dermis, to their neighboring cells in the dermis, but also to the epidermal compartments (Fig. S3A, B). Single-cell transcriptomics further showed enrichment of *LCK* expression in MF and with high presence in proliferating CD4+ T cells and T cells double-negative for CD4 and CD8 (Fig. 4A, B). In contrast to *LCK*, *HOMER1* was expressed at markedly lower levels, predominantly in keratinocytes with minor presence in stromal compartments in eczema and psoriasis and was nearly absent in MF (Fig. 4A, B). Together, these findings anchor the diagnostic performance of the classifier in the cellular and spatial distribution of its component genes.

**Fig. 4.**
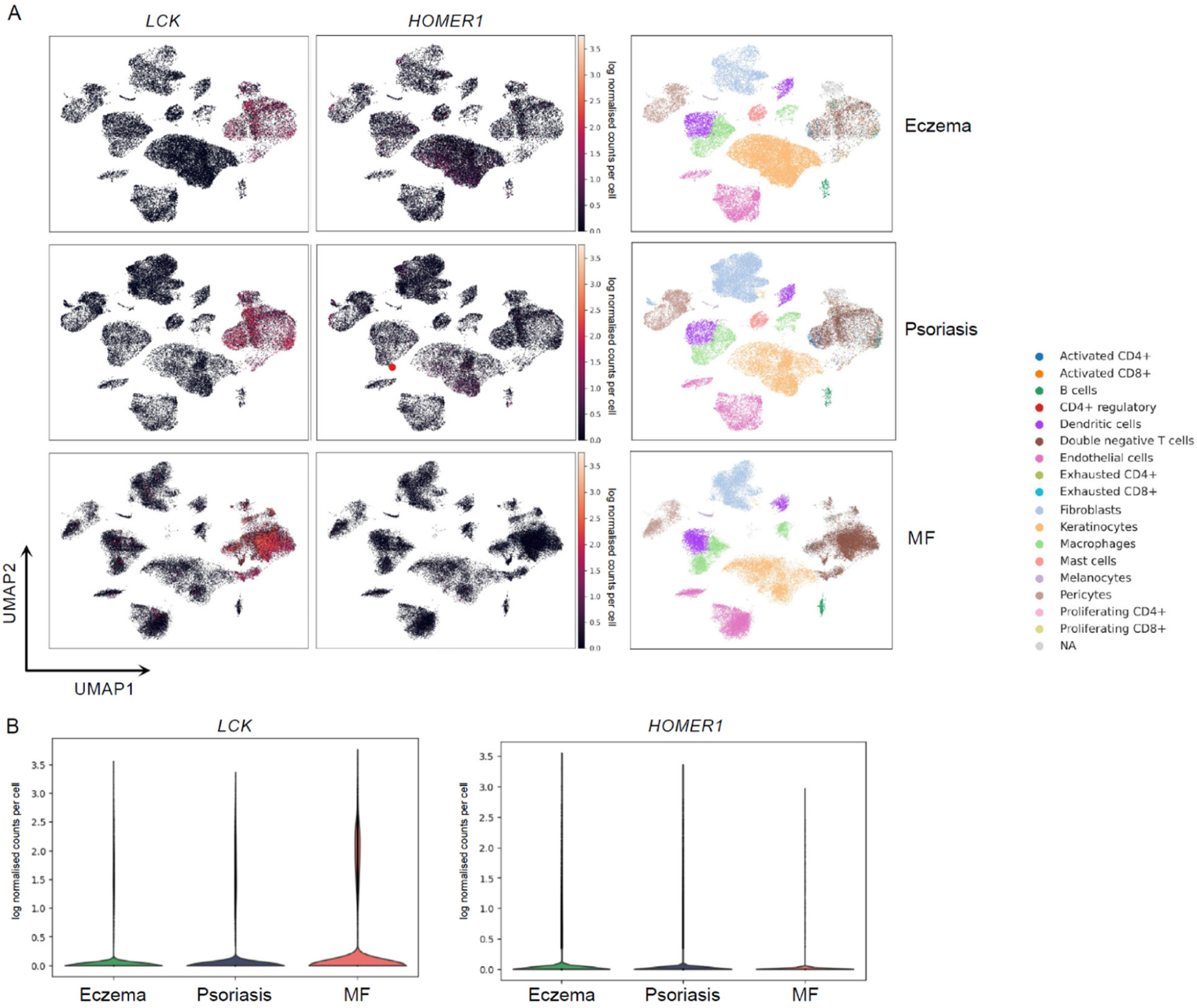
Cellular expression of LCK and HOMER1 in eczema, psoriasis and MF. **(A)** Expression of LCK and HOMER1 in single cell datasets of eczema, psoriasis and MF. Cellular subsets have been defined according to Table S5. Shown are LCK and HOMER1 log normalized counts per cell. **(B)** Violin plots summarizing log-normalized expression of LCK and HOMER1 over all single cell subsets.

### Robust validation of the two-gene classifier across seven cohorts

To assess the robustness of the two-gene classifier, we evaluated its performance in seven independent validation cohorts (VD1–VD7), comprising in total 47 MF, 58 psoriasis/eczema samples and 4 “parapsoriasis” cases (Fig. 5A, Table S1). In cohort VD1 3/28 samples were misclassified and two fell into the rejection interval (Fig. 5B). In VD2, 1/17 samples was misassigned and one eczema/psoriasis samples was rejected (Fig. 5C); notably, this rejected sample belonged to a patient that was later diagnosed with MF. In VD3, none of the 11 samples were misclassified, though two samples from 2024 and 2025 assigned to the diagnosis of eczema/psoriasis fell into the rejection interval indicating a tendency towards MF; both samples belong to two patients that are currently followed for suspected T cell lymphoma, with TCR monoclonality detected in one patient (Fig. 5D). In VD4, all patients were correctly classified (Fig. 5E), while in VD5, several misclassifications occurred, yet clinical follow-up was unavailable for four patients; one patient of these with three samples was consistently classified as MF despite a current diagnosis of eczema and is being followed up (Fig. 5F). In cohort VD6, two as eczema/psoriasis misclassified samples corresponded to one patient with clinically very stable comedogenic folliculotropic MF, and two other misclassified eczema/psoriasis samples represent unilesional, clinically stable folliculotropic MF (Fig. 5G). In VD7, 1/9 cases was misclassified as MF and two were rejected, with no follow-up available (Fig. 5H). Despite these diverse clinical scenarios, the classifier consistently assigned high MF scores to MF cases, while retaining strong specificity against inflammatory controls.

**Fig. 5.**
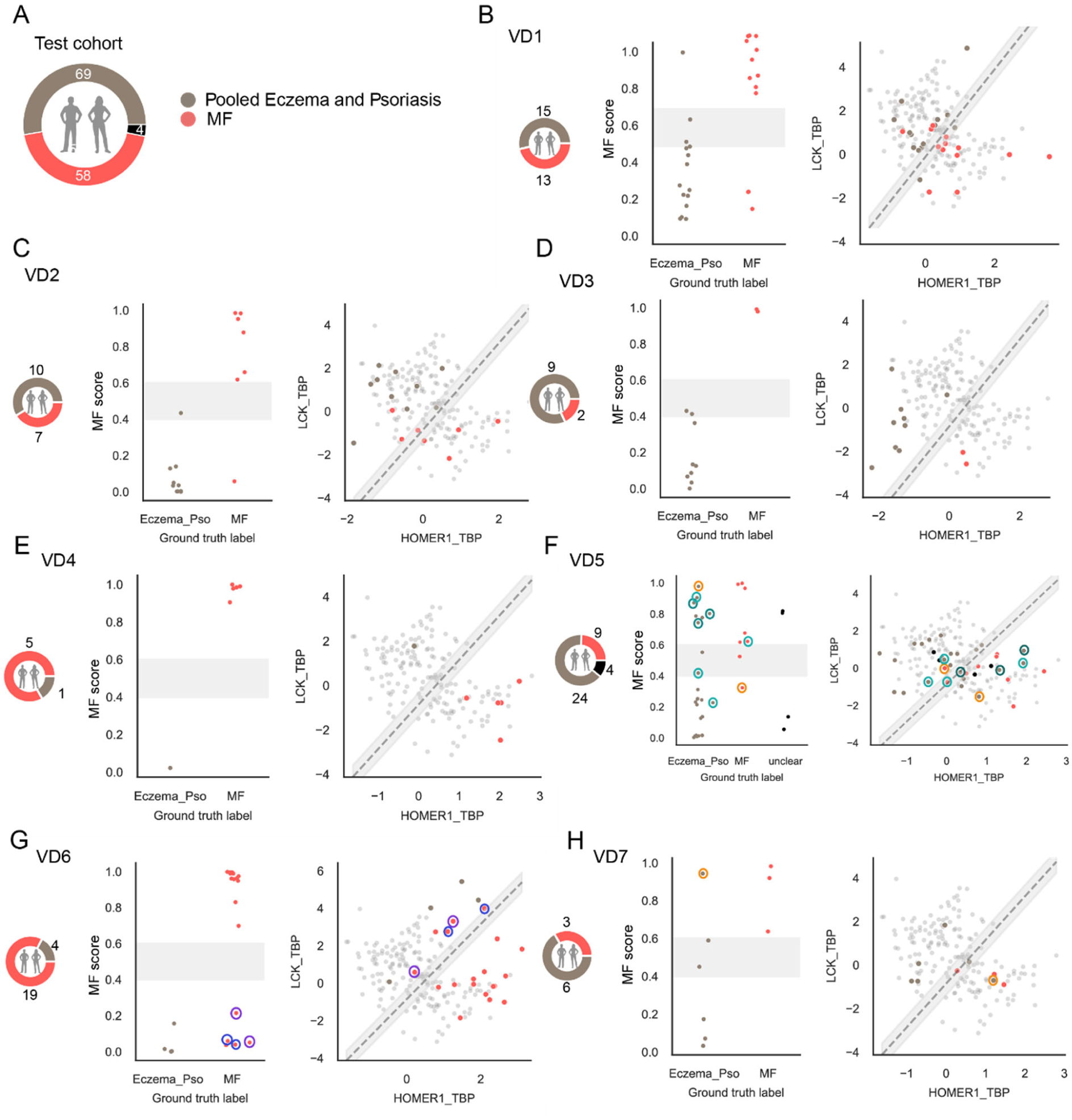
Validation of the two-gene classifier in independent test cohorts (VD1–VD7) **(A)** Summary of all validation cohorts showing the number of MF (red), pooled eczema/psoriasis (brown) and parapsoriasis (black) patients included. **(B–H)** Classifier performance in the individual cohorts VD1– VD7. Left panels display the MF score for each sample against the ground truth label; right panels show LCK vs. HOMER1 expression relative to TBP. Encircled symbols denote special case annotations: **orange** = no clinical follow-up available; **light turquoise** = samples from the same patient with follow-up scheduled; **dark turquoise** = samples from another patient with follow-up scheduled; **blue** = very stable comedogenic folliculotropic MF; **violet** = unilesional, clinically stable folliculotropic MF.

### Longitudinal cases show that the classifier detects MF in routine biopsies earlier than histopathology

To assess its potential for early detection, we applied the MF classifier to sequential biopsies from patients who showed initial clinical pictures of eczematous lesions or lesions of suspected MF and were only later clinically and histologically confirmed with MF. In all example patients shown (Fig. 6, Fig. S4), the classifier identified MF in the initial lesions, while histology only showed features of eczematous disease or an incomplete picture of MF. Patient 1, a male in his late seventies, presented with pruritic, erythematous patches initially interpreted as eczema (Fig. 6A). Histopathology revealed a band-like lymphocytic infiltrate with focal epidermotropism, CD4⁺ predominance over CD8⁺ cells, and few CD20⁺ or CD30⁺ cells, consistent with parapsoriasis, but not diagnostic for cutaneous T-cell lymphoma. At this stage the MF classifier indicated MF with a score of 0.63 (Fig. 6A). After one year, both clinical appearance and histology remained stable and were interpreted as chronic dermatitis Also the MF classifier score remained stable at 0.63 (Fig. 6B). Another year later, repeat histology demonstrated a denser, epidermotropic CD4⁺-predominant infiltrate with a CD4:CD8 ratio of approximately 4:1 and focal alignment of intraepidermal lymphocytes, indicating progression toward early mycosis fungoides, which became clinically apparent another two years later and is reflected by a MF score of 0.81 (Fig. 6C, Fig. S4A). Patient 2, a male patient in hist late sixties, presented with erythematous, pruritic plaques on the arms, upper legs, and axillae (Fig. 6D and E). Histology was consistent with acute/subacute eczema with differential diagnosis of prebullous stage of bullous pemphigoid, yet the classifier already indicated MF with scores of 0.82 and 0.94. Two years later, the patient developed worsening itch and skin involvement, and histology showed early features suggestive of MF (Fig. 6F; classifier score: 0.61). Another two years later, biopsy revealed a clear histological picture of MF (Fig. 6G), with the classifier again positive for MF (classifier score: 0.81). Additional cases of this kind are illustrated in the Supplemental Figure S4 highlighting the classifier’s power to identify MF at an earlier stage than conventional histopathology, clearing up discrepancies in clinical-histological correlation and potentially reducing diagnostic delay and guiding timely treatment decisions.

**Fig. 6.**
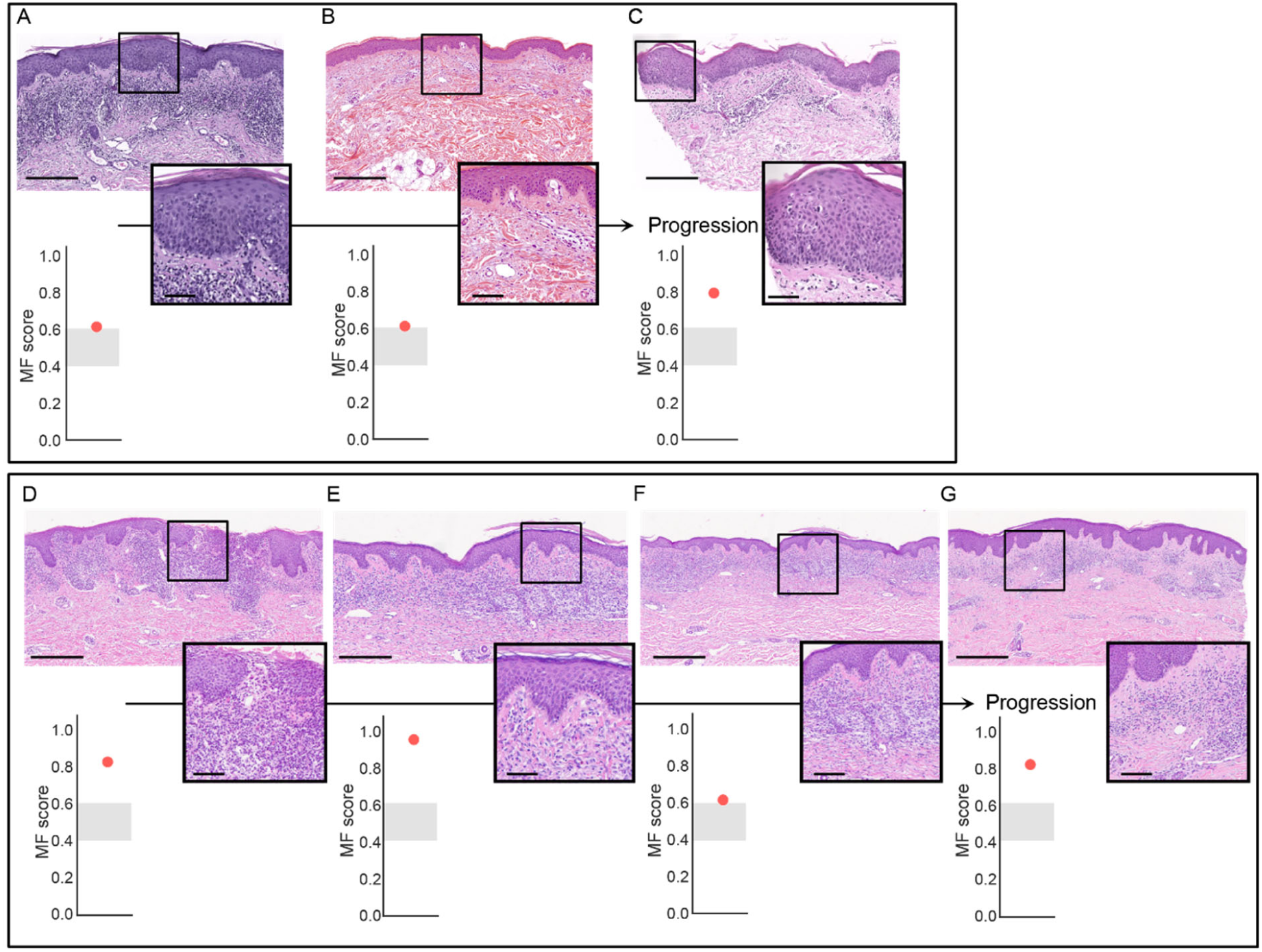
The MF Classifier detects early MF and is superior to histological diagnosis. (**A-C**) A patient in his late seventies presenting with eczematous lesions and a band-like lymphocytic infiltrate with focal epidermotropism (MF score 0.63) (A). After one year, clinical and histological findings as well as MF score remained stable (B); two years later, histology revealed an epidermotropic CD4⁺-predominant infiltrate and “wiry-collagen” in the upper dermis indicating progression toward early mycosis fungoides and accompanied by an increased MF score of 0.81; MF became clinically evident another two years later (C). (**D-G**) Clinical and histological diagnosis of a male in his late sixties with acute/subacute eczema with differential diagnosis of prebullous pemphigoid (D, E), early features of MF two years later (F), and another two years later a clear histological picture of MF (G). Again, the MF classifier identified all lesions as MF. Scale bars = 250 µm (full) and 50 µm (outtake). Clinical images of patient cases available upon request from the corresponding author.

## DISCUSSION

Early diagnosis of MF remains a major clinical and/or histopathologic challenge, as early lesions often mimic benign inflammatory dermatoses and are difficult to resolve by clinical picture and histology alone. By integrating transcriptomic profiling with a robust biomarker discovery framework, SAFARI, we developed a two-gene expression classifier that reliably separated MF from psoriasis and eczema. Validation across independent cohorts and routine FFPE samples underscored the translational potential of this tool to close a critical diagnostic gap.

SAFARI identified seven candidate biomarkers with strong discriminatory power between MF and pooled eczema/psoriasis samples. Among these, *RNF213* was notable, as Chang et al. also described it as a top tumor-defining gene in neoplastic T cells of mycosis fungoides - an encouraging overlap given the molecular heterogeneity of this disease (*30*). The strongest discriminatory signals, however, were observed for *LCK* and *HOMER1*, two genes not yet well characterized in the context of skin inflammation or cutaneous malignancies. *LCK*, a member of the Src family of non-receptor tyrosine kinases, is a central regulator of T cell receptor (TCR) signaling and immune cell activation. Upon antigen recognition, LCK phosphorylates the TCR, ZAP-70, and LAT, thereby initiating downstream signaling, but its activity also occurs independently of antigen presentation, particularly in CD8+ cells (*31*). Our pathway analyses highlight a specific role for LCK in MF, where upregulation of the ZAP-70 pathway points to aberrant TCR signaling as a potential disease-driving mechanism.

As a proto-oncogene, LCK has been implicated in tumorigenesis across several malignancies, including colorectal cancer, chronic lymphocytic leukemia, and thymomas. Thus, LCK has emerged as a therapeutic target, with several preclinical strategies under development. Dasatinib-based Proteolysis Targeting Chimera (PROTACs) degrade LCK in T-ALL models (*32*), while the selective inhibitor NTRC 0652-0 reduced YAP signaling and tumor growth in cholangiocarcinoma (*33*). Additional approaches include novel small-molecule inhibitors identified by virtual screening and molecular glue degraders that overcome resistance in T-ALL (*34*). These advances highlight the druggability of LCK, yet its function is context dependent, varying with T cell subset, age, co-receptor engagement, or viral infection (*31*). This is exemplified in skin disease: In the imiquimod-mouse model of psoriatic inflammation inhibition of LCK signaling improved clinical features, reducing inflammatory cytokines in CD4+ T cells (*35*), whereas in melanoma, higher LCK expression was correlated with greater amount of tumor-infiltrating lymphocytes and thus linked to better prognosis/staging (*36*). In MF, by contrast, malignant T cells are the primary cause of tumor and *LCK* upregulation likely reflects aberrant T cell signaling intrinsic to the tumor rather than the beneficial anti-tumor activity observed in melanoma. While *LCK* is an established oncogenic driver with emerging therapeutic inhibitors, the role of *HOMER1* is less studied. Homer-1 is a scaffolding protein of the Homer family, mainly expressed in the nervous system and at lower levels in peripheral tissues such as heart and muscle, where it contributes to synaptic plasticity and signal transduction at postsynaptic sites (*37*). However, it has not yet been described in the context of skin tumor and inflammatory skin diseases. In colorectal cancer as well as in intrahepatic cholangiocarcinoma *HOMER1* overexpression was associated with proliferation, migration and poorer survival (*38, 39*), whereas in hepatocellular carcinoma (HCC) it was downregulated and served as a highly sensitive diagnostic marker (*40*). Similar to MF, most HCC cases are diagnosed at an advanced stage due to the lack of reliable early diagnostic markers, highlighting the potential of *HOMER1* in early disease detection.

We not only developed and validated a molecular classifier that separates MF from psoriasis and eczema in routine FFPE samples. We also confirmed the classifier’s capability to identify MF earlier than standard methods highlighting its potential to reduce diagnostic delay. A representative sample of routine data from statutory health insurance funds (n = 4 million) presented by Zech at al. at the 34th German Skin Cancer Congress of the Working Group on Dermatological Oncology (ADO) 2024) showed that 35% of patients diagnosed with MF or Sézary syndrome (SS) had previously received a coding for atopic dermatitis or psoriasis (*41*). This is in accordance with a recent study from Beatty et al. confirming that 82% of surveyed dermatologists report the concern that diagnosis of early-stage MF is delayed due to misclassification as benign inflammatory dermatoses (*42*).

Today, MF is diagnosed according to the EORTC-ISCL criteria which integrate clinical presentation, histopathology including lymphoid atypia and epidermotropism, and immunophenotyping. TCR gene rearrangement is so far the only recommended molecular support (*43*). However, its use remains limited as only 2.3% of dermatologists request TCR clonality assays as part of the initial workup, and only 3.7% of pathologists rely on it for diagnosing cutaneous lymphoproliferative disorders (*6*). This likely reflects the modest sensitivity and poor specificity of this analysis with monoclonality detected in only 53 % of early stage lymphoma and monoclonality observed in 49% of benign inflammatory dermatoses (*6, 7*).

Our two-gene expression classifier could be further strengthened by integration with AI-based image analysis. Recent work demonstrated that convolutional neural networks trained on multimodal data - including clinical information, photography, and dermatoscopy - achieved higher diagnostic accuracy for distinguishing MF from inflammatory dermatoses than experienced dermatologists alone, with performance further improved when AI outputs were combined with physician assessment (*44*). Such multimodal strategies parallel the concept of the Merlin test in melanoma, which integrates molecular analysis with clinical parameters to guide decision making (*45*). Together, these approaches suggest that coupling molecular classifiers with AI-assisted imaging could provide a powerful framework for earlier, more accurate, and clinically actionable diagnosis of MF (*46*).

Beyond early diagnosis, our highly sensitive molecular classifier may also help address unresolved clinical questions where routine diagnostics fall short. For example, it remains unclear whether biologics approved for inflammatory dermatoses can unmask latent MF or promote its progression (*47*). An additional advantage of our classifier is its compatibility with archived FFPE material, allowing retrospective molecular analysis even after for example biologic therapy has been initiated. Similarly, novel agents such as mogamulizumab can cause treatment-related rashes with heterogeneous clinical presentations that are difficult to distinguish from disease progression, both clinically and on routine histologic examination (*48*). In such contexts, a two-gene expression classifier could provide an objective molecular readout to support clinical decision making where conventional histology and immunopathology are inconclusive despite close clinicopathologic correlation.

## MATERIALS AND METHODS

### Patient cohorts

The overall study included 542 samples, thereof 150 MF samples, 180 eczema samples, 168 psoriasis samples, 39 samples with overlapping eczema/psoriasis phenotype (EPV) and 5 parapsoriasis cases. Samples were divided into three independent cohorts (Table S1): ***1)*** The discovery cohort (DI) comprising bulk RNA-sequenced biopsies from Munich and Lausanne (MF n=19; eczema n=105; psoriasis n=112), ***2)*** the development cohorts (DE1/DE2): FFPE samples from Pilsen in DE1 (MF n=34; eczema n=12; psoriasis n=26; EPV n=33) and Freiburg in DE2 (MF n=31; eczema n=13; psoriasis n=11; EPV n=6), ***3)*** the validation cohorts (VD1-VD7): FFPE samples from Freiburg in VD1 (MF n=14; eczema n=14; psoriasis n=1), from Würzburg in VD2 (MF n=7; eczema n=6; psoriasis n=4), from Mainz MF n=2; eczema n=5; psoriasis n=4), from Göttingen (MF n=5; eczema n=1), from Athens (MF n=11; eczema n=14; psoriasis n=10, parapsoriasis n=5), from Zürich (MF n=19; eczema n=4) and from Dubai (MF n=3; eczema n=6). The study was approved by the local ethical committees (University of Freiburg, 25-1087-S1-AV; Technical University Munich, 166/21; University Hospital Pilsen, 123/25; University of Lausanne, CER-VD 2021-00878; University of Göttingen, 6/5/25 Ü). Patients were diagnosed according to clinical routine, which included assessment of clinical presentation, medical history, disease course, and histopathology (including immunophenotyping and molecular workup as indicated). In cases of clearly defined MF, diagnosis was established in accordance with the EORTC-ISC criteria including immunohistochemistry and TCR rearrangement.

### Library preparation and sequencing

RNA was isolated using the miRNeasy Mini Kit (Qiagen) from skin biopsies that were stored in TissueProtect (Qiagen) according to the manufacturers protocol. The Illumina TruSeq Stranded Total RNA Kit was used to generate RNASeq libraries according to the high sample protocol. Samples were sequenced on an Illumina HiSeq4000 as paired-end sequenced with a read length of 2x 150 bp and an average output of 40 M reads per sample. The read alignment was performed using STAR aligner with the human reference genome GRCh38 (*49, 50*).

Multiple steps for quality control were applied comprising removing lowly-expressed genes, genes expressed only on the Y chromosome and samples with low quality. In detail, low-expressed genes were defined as genes with less than one transcript per million. Samples exhibiting size factors within the first 0.1 quantile were filtered out due to insufficient quality. The final count matrix comprised 523 samples with 18,341 genes.

### Normalization of RNA-sequencing data

To account for different sequencing depths across samples, RNA-seq data were normalized using the trimmed mean of M-values (TMM) method (*51*). Size factors were applied to compute log counts-per-million (CPM) expression values and scaled to follow a zero-centered Gaussian with unit variance before feeding them into any machine learning model. TMM was chosen for its robustness and demonstrated concordance with qPCR data (*52*).

### Differential gene expression analysis

Differential gene expression analysis was performed in R with edgeR (*53*), comparing each disease group with non-lesional controls to identify disease-specific genes. For each gene g and sample i, raw counts y_gi were modeled with a negative binomial generalized linear model (GLM) with log link and library-size offset:

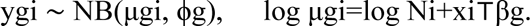

Here, μ_gi is the expected count, ϕ_g the gene-specific dispersion, N_i the total library size (offset), x_i the covariate vector (“intercept”,“condition”,“sex”,“batch”), and β_g the corresponding regression coefficients. This specification adjusts for sex and batch while estimating the effect of condition (disease vs control).

To identify genes distinguishing one disease from another, we fit contrasts on condition within the same GLM on the relevant subset; when stated, a reduced model with condition as the sole covariate was used for head-to-head discrimination. Statistical significance was assessed with edgeR’s GLM tests, and p-values were adjusted for multiple hypothesis testing using the Benjamini-Hochberg procedure. Genes were considered differentially expressed at FDR (adjusted p-value) < 0.05.

### Pathway Enrichment Analysis

Pathway enrichment analysis was conducted in R using ReactomePA (*54*). Differential expressed genes from the MF versus pooled lesional eczema and psoriasis contrast (|log2 FC| > 1; FDR < 0.05) were used as target set, and all quantified genes in the cohort defined the background universe. P-values were adjusted for multiple hypothesis testing using the Benjamini-Hochberg procedure, with FDR < 0.05 were considered significant.

### SAFARI: A computational framework for Sparse biomArkers For rARe dIseases

Gene selection in SAFARI was performed using a two-stage bootstrapping approach (nbootstrap = 200). In each iteration, a small subset of samples (nMF = nEcz|Pso = 3) was used for univariate analysis to identify genes discriminating mycosis fungoides (MF) from eczema and psoriasis by differential gene expression. Significant genes (padj < 0.05) were intersected with those differentially expressed between lesional and non-lesional samples, reducing the feature space from 18,341 to a median of 124 genes. The remaining samples were then used for multivariate feature selection via sequential forward selection using logistic regression with fourfold stacked cross-validation, L2 regularization, and inverse class weighting to correct for imbalance. Genes were iteratively added based on maximal F1-score improvement until convergence. Across 200 bootstraps, ranked gene lists were combined using RobustRank aggregation (*26*) to identify consistently informative features. Resulting p-values were corrected for the number of bootstraps (Bonferroni) and multiple testing (Benjamini–Hochberg), with genes showing adjusted p < 0.05 defined as diagnostic biomarkers.

### Establishment of the classifier

#### Evaluating SAFARI with Augmented qPCR Data

SAFARI was used to derive a linear gene signature with a logistic regression model distinguishing MF from eczema and psoriasis. During training, each sample was included twice: once in its original form and once in an augmented version where the reference genes TBP and SDHAF2 were perturbed by random noise sampled from a Gaussian distribution (mean = 0, s.d. = 0.5). This augmentation simulates measurement variability of up to ±1 in the reference genes, increasing data diversity, improving model robustness, and enhancing generalization to unseen samples.

### SAFARI: Model Comparison and Calibration on qPCR Data

Three logistic regression models were compared: (a) a baseline model using default scikit-learn parameters, (b) a model with hyperparameters optimized for the F1-score, and (c) a model with both hyperparameters and the decision threshold optimized for the F1-score (Table S2). Model performance was evaluated using 100 repetitions of 4-fold stacked cross-validation, stratified by disease and cohort to ensure disease- and cohort-independent learning (Table S3). Calibration quality was assessed using the Expected Calibration Error (ECE), defined as the weighted average of the absolute difference between predicted confidence and observed accuracy across probability bins (51), where M denotes the number of bins dividing the probability space [0,1], and B_m represents the set of samples whose predicted scores fall within bin m (Fig. S2 A) (*55*):

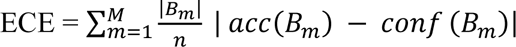

### SAFARI: Calibration Refinement and Uncertainty Handling on qPCR Data

To further improve calibration, Platt scaling (*56*) was applied using scikit-learn’s CalibratedClassifierCV function (Fig. S2A). A rejection area was introduced for predictions with probabilities between 0.4 and 0.6 to exclude uncertain classifications, balancing the fraction of rejected samples with gains in sensitivity and specificity (Fig. S2). Final predictions for the validation cohort were generated using stacked fourfold cross-validation with Platt scaling in the inner loop, whereas predictions for seven independent test cohorts were produced after model training on the full validation set with cross-validated Platt scaling to yield unbiased probability estimates. Probabilities for predictions are shown as scores and can be interpreted as follows (Table 1):

**Table 1:**
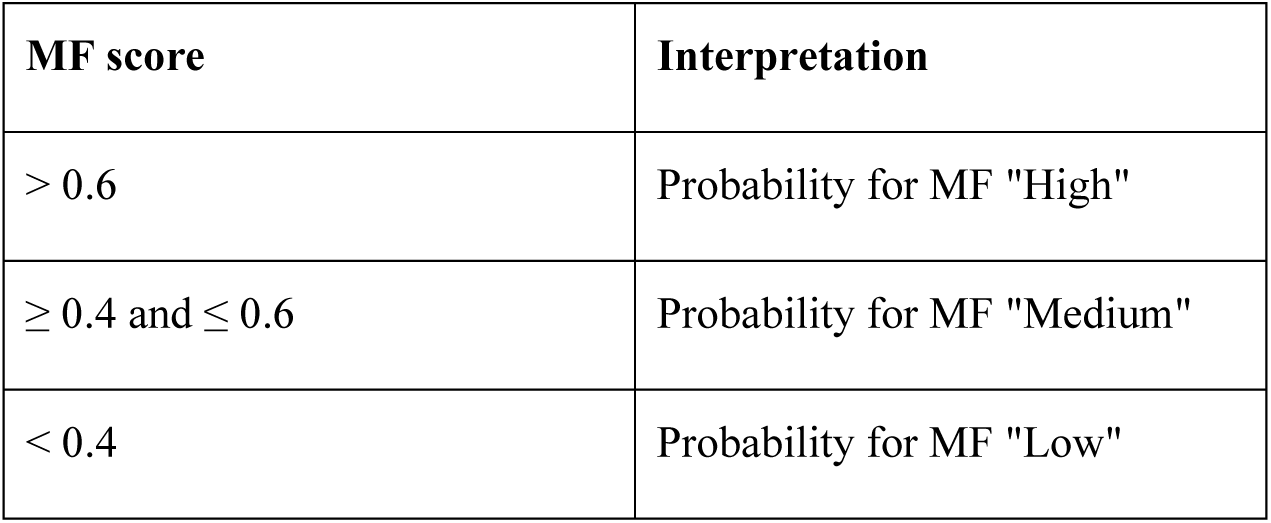
Definition of MF score.

| MF score | Interpretation |
| --- | --- |
| $> 0.6$ | Probability for MF "High" |
| $\geq 0.4$ and $\leq 0.6$ | Probability for MF "Medium" |
| $< 0.4$ | Probability for MF "Low" |

### Gene expression analysis in FFPE samples

RNA was isolated from ten sections à 10 µm of each FFPE sample using the miRNeasy FFPE Kit (Qiagen) and reversely transcribed into cDNA with the High Capacity cDNA Reverse Transcription Kit (Applied Biosystems) according to the manufacturers’ instructions. Gene expression was measured by quantitative real-time PCR using the Fast Start Universal SYBR Green Master (Rox) (Roche) system, 20 ng cDNA as input and the following target-specific primers: TBP_fw: ATGGATCAGAACAACAGCCTGC, TBP_rev: CATTGGACTAAGATAGGGATTCCG, SDHAF2_fw: CGGTGTCTACAGTGTTCTCGAC, SDHAF2_rev: GATGTCACACTGAGCAAAGGAG, HOMER1_fw: TCCGTAGCATAAGAGCTGAAAC, HOMER1_rev: TGAAGATAGGTTGTTCCCCCA, RNF213_fw: ACCGTCCAAGAAGGAGCTA, RNF213_rev: TGGCATTAGCAGCTTCCCA, ZC3H12D_fw: TTCTCTGCGACCCATAGTGA, ZC3H12D_rev: TCTTTATTTCCATGGCTCATCGC, LCK_fw: CAGACTTTGGCCTAGCACGC, LCK_rev: AATGGGAAACTTGGCCCCCTC, BTN3A1_fw: GCTGAAGGTTGCAGACGGAG, BTN3A1_rev: AGGATACACCCTCCCCAGAG, SERBP1_fw: AGCAGGACCGACAAGTCAA, SERBP1_rev: TGGCATCCAGTTAAGCCAGAG. All measurements were performed in duplicates, and samples were excluded from further analysis if the standard deviation was higher than 0.5, or if any C_T_ value was above 36. The doublets were aggregated by computing the geometric mean across the doublets. The ΔC_T_ values were calculated by subtracting the Ct values of the housekeeper genes TBP and SDHAF, respectively.

### Single-cell analysis

Two single-cell datasets (*27, 28*) were aggregated and processed in Python using scanpy (*57*), combining 7 eczema, 11 psoriasis, and 7 patch-stage MF patients, respectively. Before integration, the single-cell datasets underwent the same quality control. Cells were removed that had fewer than 500 expressed genes, a total UMI count below 600 or above 25,000, or a mitochondrial read fraction exceeding 25% of the total counts. Genes that were not expressed in at least 3 cells were removed. Doublets were filtered out using the scrublet library (*58*), with a separate run per sample. Following the QC, the datasets were combined to form an integrated matrix comprising 99,117 cells and 13,465 shared genes. The total counts per cell were normalized to a target sum of 10,000 counts to account for different sequencing depths. The normalized counts were subsequently log-transformed using log(X+1) to stabilize gene expression variance. The top 4000 highly variable genes were selected and used for follow-up dimensional reduction. The Harmony algorithm (*59*) was used to correct sample-specific batch effects. The Leiden clustering (*60*) with resolution 1 was applied on the batch-corrected neighborhood graph to annotate cells with their respective cell type. The cell markers used for this analysis are shown in Table S5.

## Data Availability

All data produced in the present study are available upon reasonable request to the authors. The code can be found here: https://github.com/MendenLab/SAFARI

## List of Supplementary Materials

Figure S1: Training and Test data for the two-gene classifier in the two independent development cohorts

Figure S2: Calibration and rejection analysis of the MF classifier.

Figure S3: Spatial and single cell expression of *LCK* and *HOMER1*.

Figure S4: The MF classifier identifies early MF lesions.

Table S1: Patient demographics

Table S2: Gridsearch hyperparameter space

Table S3: Evaluation of three different logistic regression models across 100 stacked 4-fold cross-validation runs with mean and standard deviation.

Table S4: Evaluation of the model performance predicting eczema and psoriasis in isolation across 100 repetitions with 4-fold stacked cross-validation with mean and standard deviation.

Table S5: Marker genes for the respective cell type used for single-cell annotation.

Table S6: Model performance on the discovery cohort using different models within the SAFARI framework and using the extracted biomarkers for evaluating their performance with 100 repeated 4-fold stacked cross-validation.

## Acknowledgments

MM is supported by the Helmholtz Association under the joint research school “Munich School for Data Science – MUDS”. We thank Dr. Francesco Paolo Casale for his feedback and Ann-Kathrin Fritz for her technical support.

## Author contributions

Conceptualization: KE, SE, MPM, NGS

Methodology: MM, NL, MPM, KE, SE, NGS

Investigation: MM, NL, AS, KTH, CH, SR, DK, MW, CM, BWB, YG, REB, EP, MCS, CM, WK, EG, KE, MPM, SE, NGS

Visualization: MM, SE, NGS

Project administration: MPM, SE, NGS

Supervision: MPM, SE, NGS

Writing – original draft: MM, SE, NGS

Writing – review & editing: MM, NL, AS, KTH, CH, SR, DK, MW, CM, BWB, YG, REB, EP, MCS, CM, WK, EG, KE, MPM, SE, NGS

