## Supplementary Material for "LCK and HOMER1 gene expression-based classifier distinguishes early mycosis fungoides from eczema and psoriasis"

### Supplementary Methods

#### ***Model selection within SAFARI***

SAFARI is a model-agnostic framework, that enables users to plug in any model during the forward feature selection step. In our analysis, we compared logistic regression, support-vector machine (SVM), and an xgboost model. The gene signatures derived from logistic regression and SVM are highly concordant and show superior performance compared to xgboost (Table S6).

#### ***Spatial transcriptomics analysis***

Two spatial transcriptomics datasets (*1, 2*) were processed to analyze the spatial contexts in which the two genes are expressed. The MF dataset comprises 15 early-stage MF patients, 14 with stage I MF and 1 with IIA MF. Of these, 9 patients remained in early stages (<IIB) during follow-up, while the other 6 progressed to advanced stages (>=IIB). The R library GeomxTools (*3*)was used for its processing. For quality control, the following guidelines were implemented to remove low-quality segments: a minimum of 1000 reads, 80% reads trimmed, 80% reads stitched, 80% reads aligned, 90% sequencing saturation, 2 negative control counts, 12000 maximum counts in a no template control well, 50 minimum nuclei estimated, and 800 minimum segment area. The spatial data for psoriasis and eczema, comprising 9 patients per disease, were processed as described in the original publication. The upper two epidermis layers (“upper” and “middle”) were aggregated into a single layer (“epidermis”) to match the granularity of the MF dataset.

### Supplementary Figures

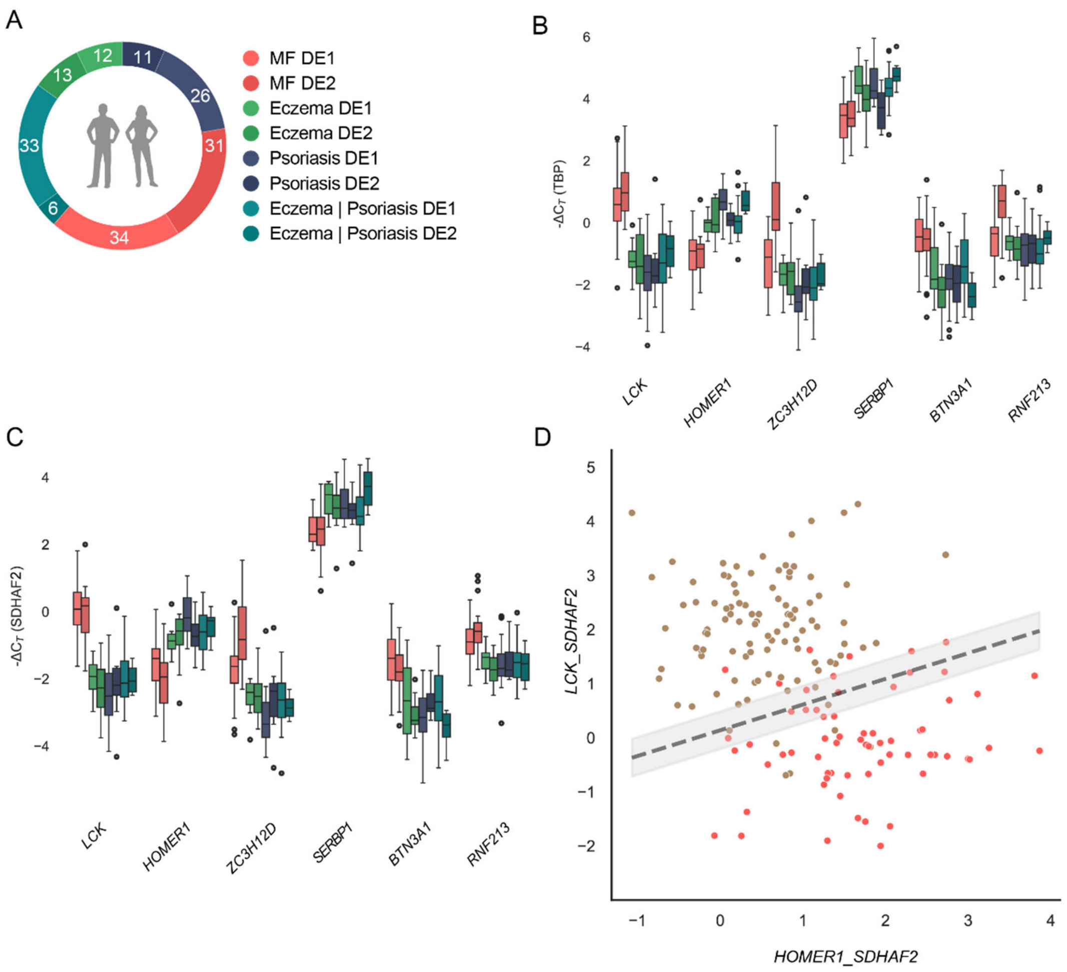


**Figure S1. Training and Test data for the two-gene classifier in the two independent development cohorts. (A)** Sample composition of the two development cohorts (DE1 and DE2) comprising MF, eczema, psoriasis and psoriasis/eczema mixed phenotype (EPV) cases. **(B, C)** qPCR expression of candidate biomarkers normalized to *TBP* (C) or *SDHAF2* (D), where eczema, psoriasis, and EPV are analyzed separately against MF, demonstrating consistent discriminatory performance across both development cohorts. **(D)** Distribution of samples in *LCK* - *HOMER1*– expression space confirms clear separation of MF from pooled eczema, psoriasis and EPV samples. **(E)** Classifier probabilities for MF show robust separation from each inflammatory subgroup, indicating that EVP, eczema and psoriasis behave similarly whether analyzed individually or pooled. EPV = eczema-psoriasis overlap variants

A

Expected calibration errors across models

| Model | Expected calibration error |
|---|---|
| Model A | 0.056 |
| Model A calibrated | **0.051** |
| Model B | 0.143 |
| Model B calibrated | 0.089 |

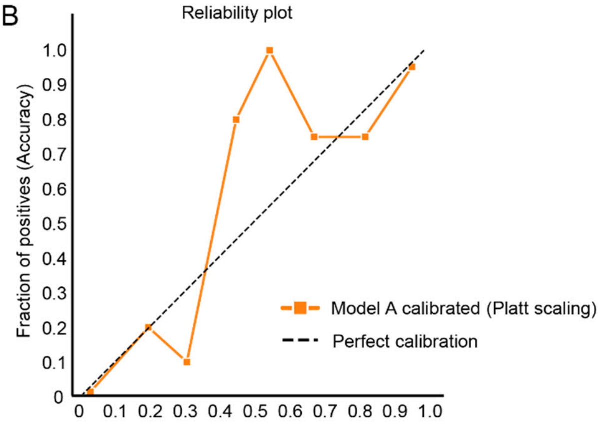


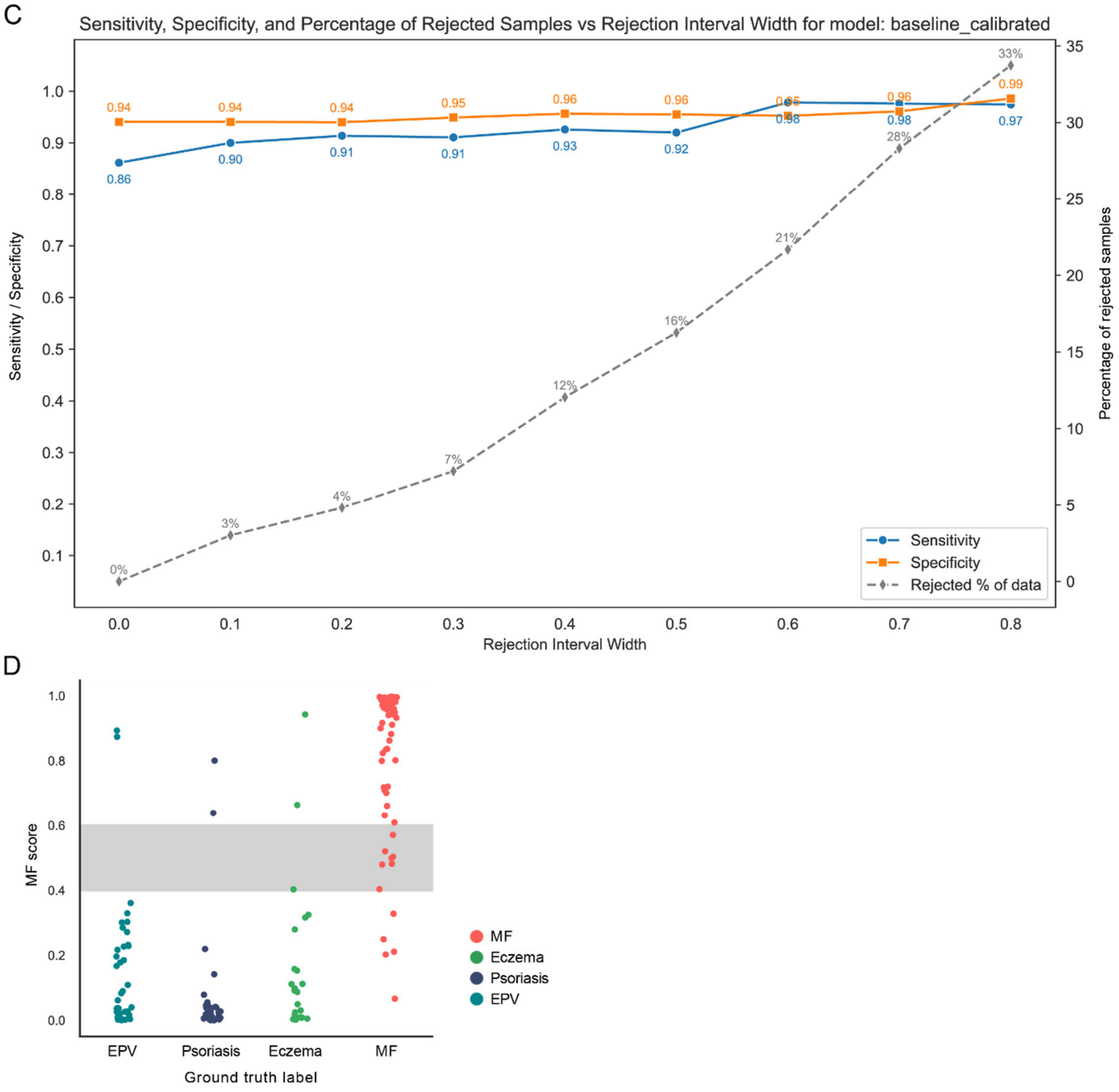

**Figure S2.** Calibration and rejection analysis of the MF classifier. **(A)** ECE for model A (logistic regression with default parameters) and model B (logistic regression optimized for F1-score via grid search). Calibration was evaluated using M = 8 bins, ensuring an average of 20 samples per bin. Model A consistently demonstrated lower ECE (better calibration) than model B, both before and after Platt scaling. **(B)** Reliability plot comparing the predicted probability (x-axis) against the true fraction of positives (observed accuracy) on the y-axis. Model A after Platt scaling demonstrates excellent calibration in the lower and upper range of probabilities, where the curve closely aligns with the perfect calibration line (grey diagonal). The model shows varying confidence with under- and overconfidence in mid-range probabilities. The interval, where the model is least certain and least reliable, was designated as the rejection area for improved downstream classification, as detailed in (C). **(C)** Sensitivity (blue), specificity (orange), and percentage of rejected samples (grey dashed line) as a function of rejection interval width in the calibrated model. Setting the rejection area to 0.4 to 0.6 reduced misclassifications, leading to a specificity of 0.94 and sensitivity of 0.91 at the cost of excluding ~4% of samples. **(D)** Probabilities of MF in all samples of the development cohort. ECE: expected calibration errors.

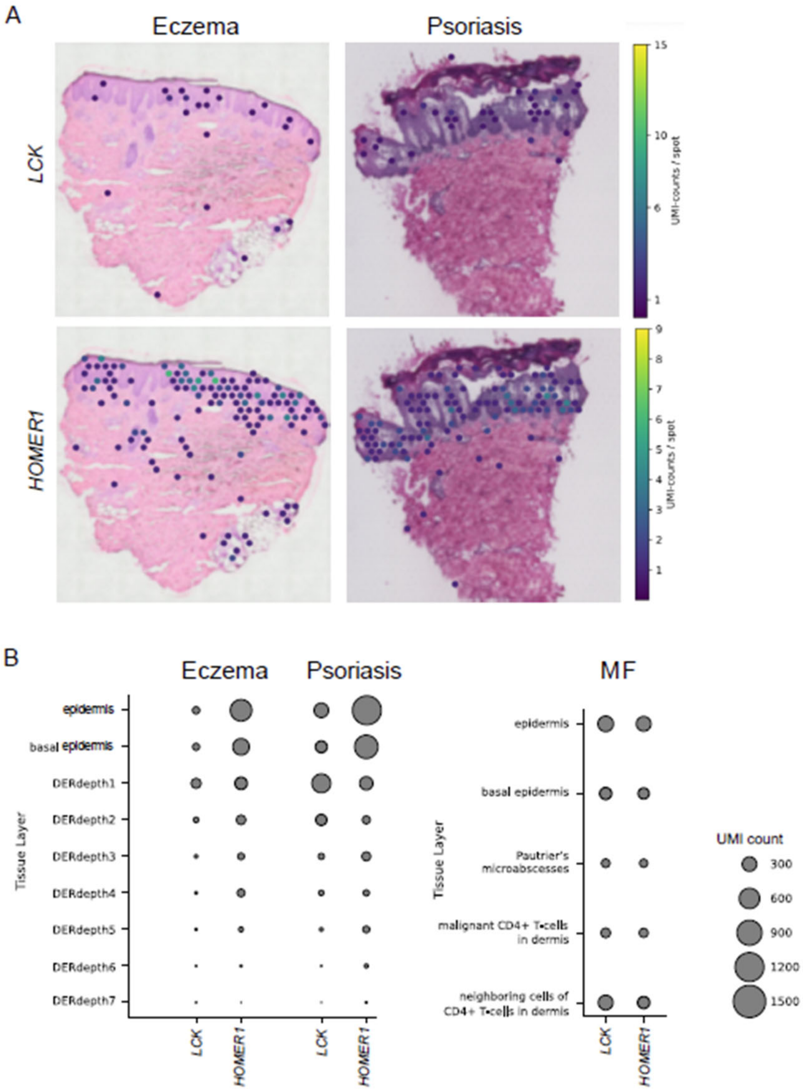


**Figure S3. Spatial and single cell expression of *LCK* and *HOMER1*.** (A) Expression of *LCK* and *HOMER1* in spatial transcriptomic maps of lesional eczema and psoriasis skin. Expression intensities are overlaid on H&E-stained histology images, highlighting disease-specific localization of *LCK* and *HOMER1*. Color code indicates transcript counts per spot. (B) Bubble plots showing spatial distribution of *LCK* and *HOMER1* across skin layers in eczema, psoriasis and MF (bubble size reflects transcript counts). DER: dermis

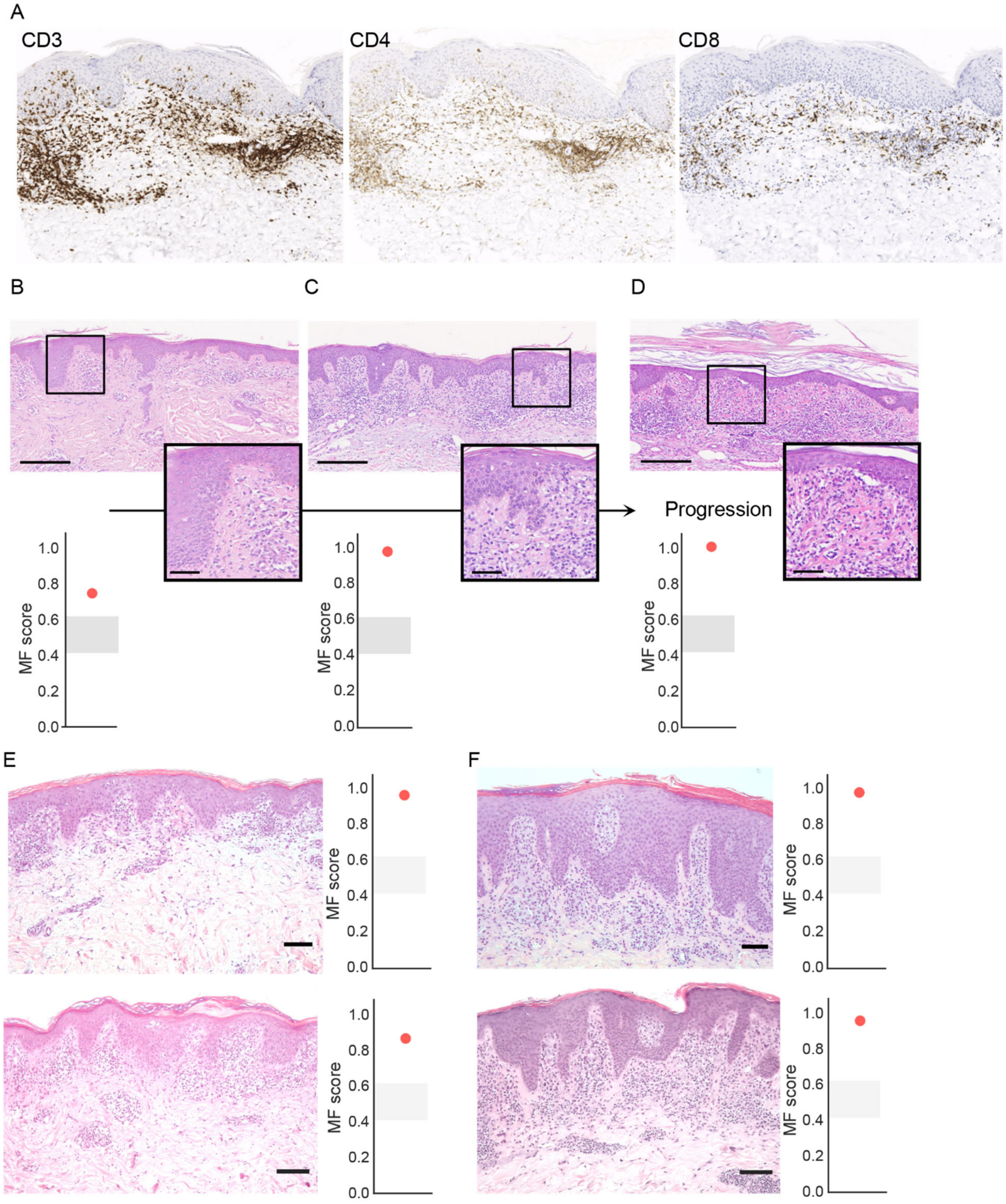


**Figure S4. The MF classifier identifies early MF lesions.** (**A**) Histological stainings for CD3, CD4 and CD8 of patient shown in Figure 6C. (**B-D**) This male patient in his seventies presented clinically with generalized xerosis, erythematous pruritic plaques, and fine lamellar scaling; histology suggested chronic eczema, yet the classifier already indicated MF (Score: 0.74) (B). Ten months later, livid scaly plaques appeared, histology shows a moderately dense, band-like lymphocytic infiltrate in the upper dermis with few eosinophils, mild spongiosis, focal parakeratosis, and discrete pigment incontinence and suspicion of early CTCL or parapsoriasis was raised, in line with an increased MF score of 0.98 (C). Weeks later, the patient presented with erythematous plaques, histology shows epidermal thinning with marked hyperparakeratosis and bacterial colonies, a superficial band-like lymphohistiocytic infiltrate with scattered eosinophils, mild epidermotropism, focal lining-up of lymphocytes, and dermal vascular dilation with erythrocyte extravasation (D). The patient is currently classified as stage IB, followed in the skin lymphoma clinic, and remains stable under phototherapy and retinoids. Scale bars (B-D) correspond to 250 µm (full) and 50 µm (outtake) (**E**) Clinical presentation of a female patient in her late forties with generalized erythematous patches and plaques; Corresponding biopsy section reveals subtle interface dermatitis-like features (middle panel), but early MF three months later (lower panel). Two-gene classifier output shows high MF probability scores for both first timepoint (0.95) and second timepoint (0.85). **(F)** A male patient in his late fifties presented with disseminated erythematous plaques on trunk and extremities. Histology was consistent with psoriasisform-lichenoid dermatitis (middle panel) and one month later lymphomatoid papulosis (lower panel). The patient is in the meantime diagnosed with MF. Again, the MF classifier shows high MF probability for both first timepoint (0.97) and second timepoint (0.94). Scale bars (E, F) correspond to 100 µm. Clinical images available upon request from the corresponding author.

### Supplementary Tables

| Cohort | Source | #Patients | #Samples | MF | Age (mean ± std) | Sex (% m) | Eczema, psoriasis | Age (mean ± std) | Sex (% m) | Para-pso-riasis | Age (mean ± std) | Sex (% m) |
|---|---|---|---|---|---|---|---|---|---|---|---|---|
| Discovery | München/ Lausanne (DI) | 233 | 236 | 19 | 72.8 ± 13.8 | 63.2 | 217 | 53.1 ± 17.4 | 67.2 | | | |
| Develop-ment | Pilsen (DE1) | 105 | 105 | 34 | 62.4 ± 17.0 | 55.9 | 71 | 47.4 ± 17.2 | 54.3 | | | |
| | Freiburg (DE2) | 50 | 61 | 31 | 63.2 ± 11.7 | 66.7 | 30 | 55.8 ± 17.6 | 63.0 | | | |
| Validation | Freiburg (VD1) | 9 | 29 | 14 | 68.4 ± 9.2 | 77.2 | 15 | 68.9 ± 9.7 | 87.50 | | | |
| | Würzburg (VD2) | 15 | 17 | 7 | 64.3 ± 7.8 | 1.0 | 10 | 53.8 ± 18.9 | 80.0 | | | |
| | Mainz (VD3) | 4 | 11 | 2 | 77 ± 0 | 0.0 | 9 | 63.0 ± 8.6 | 66.7 | | | |
| | Göttingen (VD4) | 3 | 6 | 5 | 58.3 ± 25.7 | 0.0 | 1 | 86 ± 0 | 0.00 | | | |
| | Athens (VD5) | 29 | 40 | 10 | 61.9 ± 15.8 | 37.5 | 25 | 63.0 ± 9.3 | 50.0 | 5 | 69.3 ± 7.6 | 66.7 |
| | Zürich (VD6) | 22 | 23 | 19 | 63.2 ± 16.6 | 77.8 | 4 | 64.5 ± 15.2 | 50.00 | | | |
| | Dubai (VD7) | 9 | 9 | 3 | 32.7 ± 10.1 | 33.3 | 6 | | | | | |

**Table S1. Patient demographics.** Patient cohorts included in discovery, development, and validation phases. Overview of patient cohorts with number of patients, number of samples, and distribution across diagnoses (mycosis fungoides [MF], eczema, psoriasis, and parapsoriasis). For each cohort, mean age standard deviation (std) and sex distribution (% male) are shown separately for MF and eczema/psoriasis cases.

| **Hyperparameter** | **Hyperparameter space** |
|---|---|
| Penalty | L1, L2, Elasticnet |
| C | 0.001, 0.003, 0.007, 0.019, 0.052, 0.139, 0.373, 1.0 |
| Elasticnet ratio | 0, 0.1, 0.2, 0.3, 0.4, 0.5, 0.6, 0.7, 0.8, 0.9, 1.0 |
| Class weight | Balanced, (0:1, 1:2), (0:1, 1:3), (0:1, 1:4), (0:1, 1:5), (0:1: 1:6) (0:1, 1:7) |

**Table S2. Gridsearch hyperparameter space.** Overview of the hyperparameters that were optimized with gridsearch with respect to the F1-score on the left, with the respective search space on the right. The hyperparameter names correspond to the hyperparameters for sklearn's logistic regression model.

| Model | Balanced accuracy | Sensitivity | Specificity | F1 score | ROC AUC | PR AUC |
|---|---|---|---|---|---|---|
| (A) | 91.4±0.9% | 90.2±1.6% | 92.7±0.9% | 89.5±1.1% | 96.5±0.3 | 94.7±0.4 |
| (B) | 91.7±0.8% | 91.4 ±1.4% | 92.0±1.1% | 89.7±1.0% | 95.4±1.0 | 92.5±1.9 |
| (C) | 91.1±1.2% | 91.1±1.2% | 91.7±1.7% | 89.0±1.4% | 95.4±1.0 | 92.5±1.9 |

**Table S3.** Evaluation of three different logistic regression models across 100 stacked 4-fold cross-validation runs with mean and standard deviation. Model (A) corresponds to the default sklearn logistic regression model with L2-regularization (C=1) and class imbalance accounting by setting the class_weight argument to "balanced". Model (B) is tuned using an inner 5-fold cross-validation to find the best parameters from Table S2, optimizing the F1-score. Model (C) uses two nested 5-fold cross-validations, where the inner one is used to find the best hyperparameters from Table S2, as model (B). The outer one is used to find the best decision threshold for the learned hyperparameters. All cross-validations use stratification by disease and cohort to learn disease- and cohort-independent parameters. The optimization metric is the F1 score. The maximum performance per metric across the three different models is highlighted in bold, revealing that the more complex models (B) and (C) do not outperform model (A).

| Disease | Balanced accuracy | Sensitivity | Specificity | F1 score | ROC AUC | PR AUC |
|---|---|---|---|---|---|---|
| Eczema | 90.1±1.4% | 90.2±1.6% | 90.0±2.4% | 92.9±1.0% | 95.0±0.4 | 97.8±0.2 |
| Psoriasis | 92.3±0.8% | 90.2±1.6% | 94.4±0.6% | 93.3±0.9% | 98.4±0.2 | 99.0±0.1 |

**Table S4.** Evaluation of the model performance predicting eczema and psoriasis in isolation across 100 repetitions with 4-fold stacked cross-validation with mean and standard deviation. Analogously to table S3, the performance of the logistic regression model with L2 regularization (C=1) and balanced class weighting was evaluated using samples from psoriasis, eczema, and MF in the training folds, with predictions made solely on MF and eczema or psoriasis samples, respectively.

| Cell type | Marker genes |
|---|---|
| Activated CD4+ | CD3D+ CD4+ TNFRSF4+ |
| Activated CD8+ | CD3D+ CD8A+ TNFRSF4+ |
| B cells | MS4A1+ |
| CD4+ regulatory | CD3D+ CD4+ FOXP3+ |
| Dendritic cells | CD1C+ |
| Double negative T cells | CD3D+ CD4- CD8A- |
| Endothelial cells | VWF+ |
| Exhausted CD4+ | CD3D+ CD4+ CXCL13+ |
| Exhausted CD8+ | CD3D+ CD8A+ CXCL13+ |
| Fibroblasts | COL1A1+ |
| Keratinocytes | KRT1 |
| Macrophages | AIF1+ |
| Mast cells | TPSAB1+ |
| Melanocytes | PMEL+ |
| Pericytes | RGS5+ |
| Proliferating CD4+ | CD3D+ CD4+ MKI67+ |
| Proliferating CD8+ | CD3D+ CD8A+ MKI67+ |

**Table S5. Marker genes for the respective cell type used for single-cell annotation.** On the left, the cell type is shown with its respective cell markers on the right. The + indicates the marker gene must be expressed for the cells in the respective cell type, while the – indicates the absence of the respective gene in that cell type.

| Model | Number of genes | Balanced accuracy | Sensitivity | Specificity | F1 score | ROC AUC | PR AUC |
|---|---|---|---|---|---|---|---|
| Logistic regression | 7 | 90.9 ±1.0% | 87.7±1.9% | 94.2±0.9% | 70.0±2.8% | 97.2±1.3 | 99.7±1.7 |
| Support vector machine | 6 | 91.4±2.7% | 90.4±3.5% | 92.3±1.0% | 65.9±3.5% | 94.9±2.3 | 99.4±0.4 |
| XGBoost | 3 | 84.6±4.5% | 73.1±9.0% | 96.2±0.9% | 68.0±6.2% | 92.4±2.4 | 98.8±0.6 |

**Table S6. Model performance.** Performance of the model on the discovery cohort using different models within the SAFARI framework and using the extracted biomarkers for evaluating their performance with

100 repeated 4-fold stacked cross-validation. The mean and standard deviation across the 100 runs per metric are reported for each model. The models were trained using an inner 3-fold cross-validation stratified by disease to find the best hyperparameters optimizing the F1-score with gridsearch.

### *References*


1. A. Schabitz, C. Hillig, M. Mubarak, M. Jargosch, A. Farnoud, E. Scala, N. Kurzen, A. C. Pilz, N. Bhalla, J. Thomas, M. Stahle, T. Biedermann, C. B. Schmidt-Weber, F. Theis, N. Garzorz-Stark, K. Eyerich, M. P. Menden, S. Eyerich, Spatial transcriptomics landscape of lesions from non-communicable inflammatory skin diseases. *Nat Commun* **13**, 7729 (2022).
2. M. Danielsen, T. Emmanuel, M. M. Nielsen, L. M. Lindahl, M. Gluud, N. Ødum, L. Raaby, T. Steiniche, L. Iversen, R. Bech, RUNX2 as a novel biomarker for early identification of patients progressing to advanced-stage mycosis fungoides. *Frontiers in Oncology* **14**, 1421443 (2024).
3. M. Griswold, N. Ortogero, Z. Yang, R. Vitancol, D. Henderson, GeomxTools: nanoString geoMx tools. *R Package Version* **3**,  (2024).
